# INTERNATIONAL EVALUATION OF AN ARTIFICIAL INTELLIGENCE-POWERED ECG MODEL DETECTING OCCLUSION MYOCARDIAL INFARCTION

**DOI:** 10.1101/2023.04.26.23289180

**Authors:** Robert Herman, H. Pendell Meyers, Stephen W. Smith, Dario T. Bertolone, Attilio Leone, Konstantinos Bermpeis, Michele M. Viscusi, Marta Belmonte, Anthony Demolder, Vladimir Boza, Boris Vavrik, Viera Kresnakova, Andrej Iring, Michal Martonak, Jakub Bahyl, Timea Kisova, Dan Schelfaut, Marc Vanderheyden, Leor Perl, Emre K. Aslanger, Robert Hatala, Wojtek Wojakowski, Jozef Bartunek, Emanuele Barbato

**Author notes:** Address for correspondence: Robert Herman, MD Powerful Medical Bratislavská 81/37, Šamorín, Slovakia Emanuele Barbato, MD, PhD Department of Clinical and Molecular Medicine Sapienza University of Rome Via di Grottarossa 1035, 00189 Rome, Italy.

## Abstract

**Background:** One third of Non-ST-elevation myocardial infarction (NSTEMI) patients present with an acutely occluded culprit coronary artery (occlusion myocardial infarction [OMI]), which is associated with poor short and long-term outcomes due to delayed identification and consequent delayed invasive management. We sought to develop and validate a versatile artificial intelligence (AI)-model detecting OMI on single standard 12-lead electrocardiograms (ECGs) and compare its performance to existing state-of-the-art diagnostic criteria.

**Methods:** An AI model was developed using 18,616 ECGs from 10,692 unique contacts (22.9% OMI) of 10,543 patients (age 66±14 years, 65.9% males) with acute coronary syndrome (ACS) originating from an international online database and a tertiary care center. This AI model was tested on an international test set of 3,254 ECGs from 2,263 unique contacts (20% OMI) of 2,222 patients (age 62±14 years, 67% males) and compared with STEMI criteria and annotations of ECG experts in detecting OMI on 12-lead ECGs using sensitivity, specificity, predictive values and time to OMI diagnosis. OMI was based on a combination of angiographic and biomarker outcomes.

**Results:** The AI model achieved an area under the curve (AUC) of 0.941 (95% CI: 0.926-0.954) in identifying the primary outcome of OMI, with superior performance (accuracy 90.7% [95% CI: 89.5-91.9], sensitivity 82.6% [95% CI: 78.9-86.1], specificity 92.8 [95% CI: 91.5-93.9]) compared to STEMI criteria (accuracy 84.9% [95% CI: 83.5-86.3], sensitivity 34.4% [95% CI: 30.0-38.8], specificity 97.6% [95% CI: 96.8-98.2]) and similar performance compared to ECG experts (accuracy 91.2% [95% CI: 90.0-92.4], sensitivity 75.9% [95% CI: 71.9-80.0], specificity 95.0 [95% CI: 94.0-96.0]). The average time from presentation to a correct diagnosis of OMI was significantly shorter when relying on the AI model compared to STEMI criteria (2.0 vs. 4.9 hours, p<0.001).

**Conclusions:** The present novel ECG AI model demonstrates superior accuracy and earlier diagnosis of AI to detect acute OMI when compared to the STEMI criteria. Its external and international validation suggests its potential to improve ACS patient triage with timely referral for immediate revascularization.

**CLINICAL PERSPECTIVE:** *What is new?:*

- A novel artificial intelligence (AI) model detecting acute occluded coronary artery (OMI) using standard 12-lead electrocardiograms (ECGs) was developed from an international cohort.
- The OMI AI model is the first of its kind to be validated in an external international cohort of patients using an objective angiographically confirmed endpoint of OMI.
- Our study demonstrated the OMI AI models superior accuracy in identifying OMI and shorter time to correct diagnosis compared to standard of care STEMI criteria.

*What are the clinical implications?:*

- The OMI AI model has the potential to improve ACS triage and clinical decision-making by enabling timely and accurate detection of OMI in NSTEMI patients.
- The robustness and versatility of the OMI AI model indicate its potential for real-world clinical implementation in ECG devices from multiple vendors.
- Prospective studies are essential to evaluate the efficacy of the OMI AI model and its impact on patient outcomes in real-world settings.

## INTRODUCTION

Patients with an acutely occluded coronary artery (Occlusion Myocardial Infarction, or OMI) who will benefit from emergent reperfusion therapy are currently identified mainly on the basis of electrocardiographic ST-segment elevation (ST Elevation Myocardial Infarction [STEMI]), according to the universal guidelines.^1,2^ Growing evidence suggests that the current acute coronary syndrome (ACS) classification dichotomizing patients as STEMI or non-STEMI (NSTEMI) is unsatisfactory for the timely diagnosis of OMI, as also recognized by the 2022 American College of Cardiology Chest Pain Expert Consensus.^3^ On one hand, 25% to 30% of NSTEMI patients present with acute coronary occlusion with insufficient collateral circulation as discovered only on delayed coronary angiography.^4^ The delayed invasive management in these patients is associated with higher short and long-term mortality.^4,5^ On the other hand, catheterization laboratories are inappropriately activated in 15-35% of suspected STEMIs where eventually no culprit lesions or a non-ischemic etiology of ST elevation is found.^6-8^ A plethora of ECG criteria have been proposed to increase the diagnostic sensitivity for OMI as compared to the current guideline-based STEMI criteria, and to differentiate OMI from mimics.^3,5,9-15^ However, their adoption is limited due to their complexity and unclear inter-evaluator reliability.

The application of artificial intelligence (AI) to ECG waveforms has demonstrated increased diagnostic accuracy in various conditions and may offer a significant improvement in the timely detection of OMI.^16-20^ Therefore, we developed an automated deep learning AI model detecting acute OMI using only single standard 12-lead ECGs as input, and hypothesized that it would outperform the existing state-of-the-art ECG criteria for detection of acute OMI and perform equally to interpreters with special expertise in ECG OMI diagnosis in patients with suspected ACS.

## METHODS

### Study design

This is a retrospective study following three key stages: (1) development of a Powerful Medical (PM)cardio-OMI AI model for the detection of acute OMI using only 12-lead electrocardiograms as input (“derivation cohort”); (2) blinded AI model evaluation on an internal European testing dataset (“EU internal test set”); (3) blinded AI model evaluation in an independent external United States (“US external test set”). In the analysis, the term “overall test set” encompasses all ECGs that were contained in the combined EU internal test set and the US external test set. Each of these steps are described below.

### Data sources and processing

The derivation cohort included data from 12,241 patients undergoing coronary angiography and serial troponin testing between 2011 and 2021 at the Cardiovascular Centre Aalst in Belgium and an online international database of 2,368 ACS patients (see Supplemental material for detailed description). Waveform data were exported from the MUSE ECG data management system (GE Healthcare, Chicago, IL) in XML format and sampled at 500 Hz. Patients without acute symptoms compatible with ACS undergoing coronary angiogram (CAG) were identified using manual chart review and excluded. ECGs recorded more than 24 hours before CAG and all ECGs post-CAG were removed. The remaining patients and contacts retained in the final dataset were carefully split into a model development (derivation) set and an internal EU testing dataset ensuring that patients with more than one (recurrent) ACS contacts were present in only one of the sets. Time from the first ECG to intervention was recorded for all cases if the patient underwent coronary angiography. The derivation set included ECGs in the EU dataset which were classified as OMI or not OMI by interpreters with special expertise in ECG OMI diagnosis (SWS, HPM), and by outcome data (see details below under “model development”). “Not OMI” includes patients who either have no acute MI or have acute Non-Occlusion MI (Non-OMI, or NOMI). Images of ECG tracings from multiple device vendors within the online database of ACS patients were converted to digital waveforms using proprietary CE-certified PMcardio ECG digitization technology (Powerful Medical, Samorin, Slovakia). The full overview of data sources and inclusions and exclusions are available in Figure 1.

**Figure 1.**
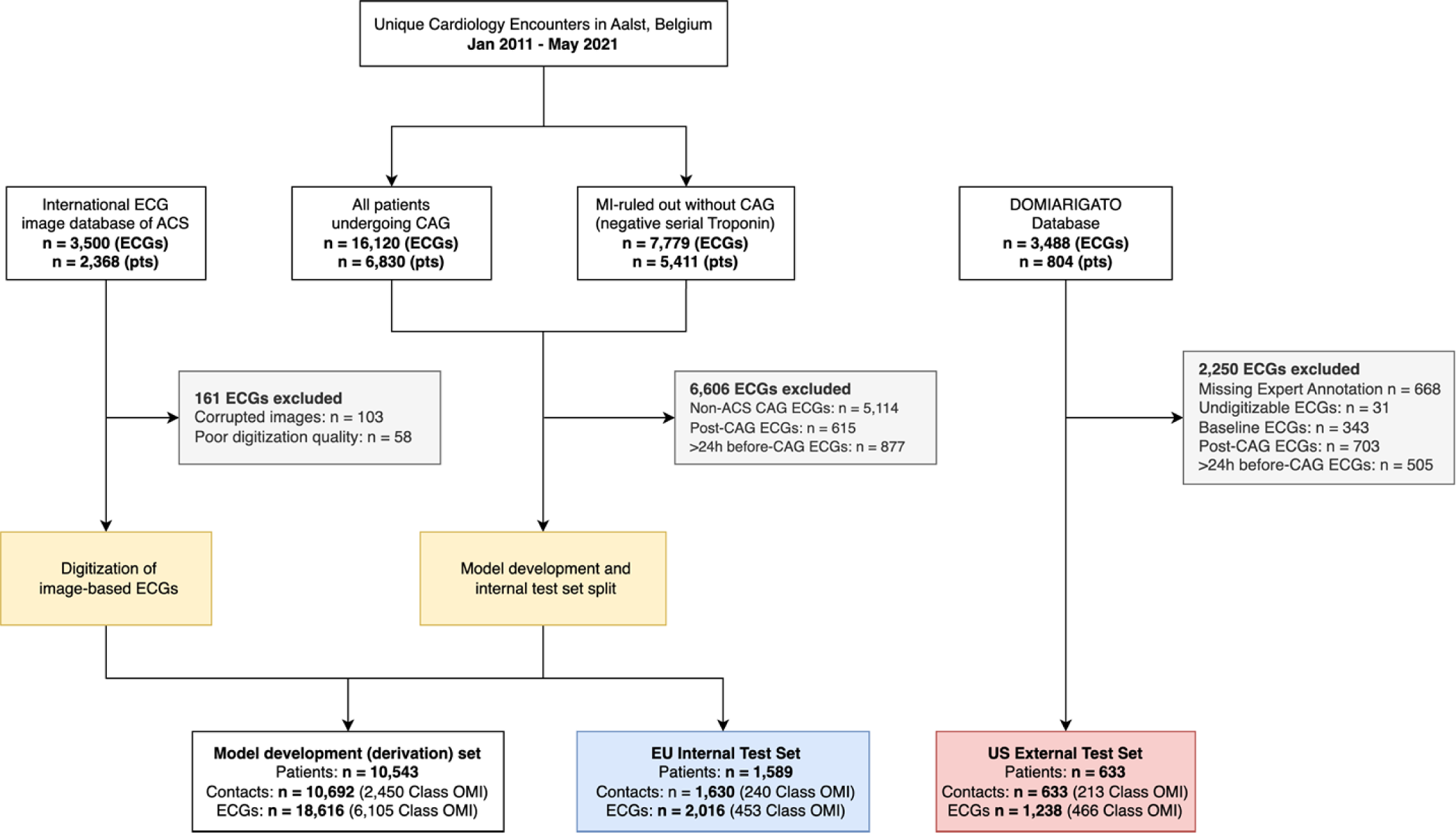
PRISMA Flow chart showing data sources and study populations. *Unique cardiology contacts identified, exclusions (in grey), and final study population split into model development (development) set (in white), EU internal test set (in blue) and US external test set (in red).* ECG, electrocardiogram; ACS, acute coronary syndrome; pts, patients; CAG, coronary angiography; MI, myocardial infarction; OMI, occlusion myocardial infarction.

### Primary and secondary outcomes

The primary outcome was the AI model’s ability to identify patients with angiographically confirmed OMI using single standard 12-lead ECGs alone. The primary definition of OMI was reproduced from previous studies^5,9,10,21-24^ and consisted of coronary angiographic evidence of an acute culprit stenosis with either a) Thrombolysis in Myocardial Infarction (TIMI) flow grade of 0-2 and any positive troponin; or b) TIMI flow grade of 3 and a very high peak troponin elevation (hs-cTnT ≥ 1,000 ng/L, cTnI of > 10.0 ng/mL, or cTnT of > 1.0 ng/mL).^25^ The primary outcome does not encompass chronic total occlusions (CTO) and all types of acute myocardial infarctions (AMI) for 2 reasons: 1) AMI that is not OMI and without persistent severe ischemia is often undetectable or nonspecific on the ECG and 2) such AMI without persistent occlusion or persistent severe ischemia does not need emergent intervention and may be diagnosed with some delay based on troponin assessment. This outcome was considered the reference standard for all analyses unless otherwise specified.

Secondary outcomes included: i) AI model performance in the subgroup analyses; ii) comparison of the AI model performance against existing criteria for detecting acute coronary occlusion (ACO) from 12-lead ECGs, iii) analysis of AI model performance using different definitions of OMI combining culprit vessel TIMI flow and peak troponin cut-offs, and iv) analysis of misclassified cases.

### OMI AI model development

Digital and digitized 12-lead ECG input data collected from sources described above was standardized into 3x4 ECG format. The model derivation set was further subdivided into a training and validation set. A deep convolutional neural network (DCNN) architecture was deployed in model development and included two key components: feature extraction and classification. The feature-extraction component, comprised of 15 Convolutional layers, was designed to extract features in a lead-specific manner. Lead-specific analysis was achieved by implementing parallel convolutional layers focused on analyzing separate leads. The second component, classification, combined all extracted features and processed them through 3 fully connected layers, interspersed with dropouts. Analysis of each lead, and integration of the knowledge gained, mimics the analytical approach of human experts to make a final diagnosis. The validation data set was used for hyperparameter refinement and threshold selection. The optimal model threshold was selected using receiver operating characteristics (ROC) curve analysis.^26^ An additional threshold was selected to match the specificity of the STEMI criteria.

### EU internal testing dataset

Independent clinical reviewers verified the angiographic data of all patients included in the EU internal testing dataset. Verification included blinded identification of culprit vessels, their visual assessment of coronary stenosis, TIMI flow, presence of sufficient collateral flow on all individual angiograms and documentation of treatment strategy. If applicable, revascularization time, defined as the duration between the first ECG and the time when a balloon was inflated or when the wire crossed the lesion, was documented.

### US external testing dataset

ECG and outcome data from the Diagnosis of Occlusion MI And Reperfusion by Interpretation of the electrocardioGram in Acute Thrombotic Occlusion (DOMI ARIGATO) database (clinical trials.gov number NCT03863327) were included in the US external testing cohort. Data collection and processing of this database is explained in detail elsewhere.^21^ Briefly, the DOMI ARIGATO database collected ECGs, laboratory and angiographic data of patients presenting with ACS to two US sites, Stony Brook University Hospital (SBUH) and Hennepin County Medical Center (HCMC). ECGs were interpreted and manually annotated by both ECG experts blinded to all clinical data other than age and sex. Baseline ECGs, post-CAG ECGs and ECGs with missing expert annotations were removed from the testing cohort.

### Benchmarking

The developed AI model was compared to three current standard criteria for detecting OMI on 12-lead ECGs: 1) physician annotation of STEMI criteria as a surrogate finding for OMI (“STEMI criteria”), 2) subjective ECG expert annotation of OMI (“ECG Experts”), and 3) a prior CE-approved AI-model trained to detect STEMI (“PMcardio-STEMI AI Model” [Powerful Medical, Samorin, Slovakia]). For criterion 1, the classification of STEMI was based on the 4^th^ universal definition of myocardial infarction.^27^ For criterion 2, independent ECG experts (SWS, HPM) with consolidated expertise in OMI detection annotated all tracings for the presence of OMI, blinded to all clinical information.^9^ For criterion 3, the prior PMcardio-STEMI AI model (trained on General Electric [GE] Marquette 12SL and/or physician annotations of STEMI criteria) was used to collect continuous predictions. All ECGs in the EU internal testing dataset, and US external testing dataset were labeled using the three methods described in this paragraph. Time to diagnose OMI was noted for each criterion by measuring the duration from the initial ECG to the accurate identification of OMI. In cases where the criterion failed to detect OMI in any ECG, the time to diagnosis was equivalent to the time to CAG.

### Statistical analyses

Statistical analysis was performed using python programming language and the following open-source libraries *tableone*, *lifelines,* and *pandas*. Continuous statistics with normal distribution were expressed as mean ± standard deviation (SD) and compared by students’ t-tests. Continuous variables with a non-normal distribution were presented as median with interquartile ranges (IQR) and reached by the Mann-Whitney-U test.^28^ If appropriate, categorical variables were reported by frequencies and percentages and compared with the Chi-square test and a Fisher’s exact test. The performance of the AI models, ECG experts, and STEMI criteria was evaluated using the following standard evaluation metrics: sensitivity, specificity, accuracy, negative predictive value (NPV), positive predictive value (PPV), Matthew’s correlation coefficient (MCC) and area under curve (AUC). For all evaluation metrics, we estimated the confidence intervals at 95% by 10,000 iterations of the bootstrap method.^29^ In the subgroup analysis, patients’ ECGs were stratified according to ECG measurement (QRS duration and heart rate) and ECG diagnostic annotations (rhythm, ventricular hypertrophy, bundle branch blocks) originating from CE-certified PMcardio AI ECG interpretation technology.

## RESULTS

### Sample characteristics

A total of 18,616 ECGs from 10,692 unique contacts (22.9% OMI) of 10,543 patients (age 66±14 years, 65.9% males) from the Cardiovascular Center in Aalst and from the international online ACS database were included in the AI model development. Sample characteristics are shown in Table 1.

**Table 1.** Sample characteristics of the model development and EU and US test sets.

| Parameter | Cat. | Model development set (n=18,616) | Overall Test Set (n=3,254) | P-value (All) | Internal EU Test Set (n=2,016) | External US Test Set (n=1,238) | P-value (Overall Test Sets) |
| --- | --- | --- | --- | --- | --- | --- | --- |
| Unique Patients, n |  | 10,543 | 2,222 |  | 1,589 | 633 |  |
| Age [years], mean (SD) |  | 66 (14.0) | 62 (14.0) | <0.001 | 63 (14.0) | 61 (14.0) | <0.001 |
| Gender, n (%) | *Female* | 3,394 (34.1) | 747 (33.0) | 0.336 | 543 (33.3) | 204 (32.2) | 0.658 |
|  | *Male* | 6,560 (65.9) | 1,516 (67.0) | 0.336 | 1,087 (66.7) | 429 (67.8) | 0.658 |
| Unique Contacts, n |  | 10,692 | 2,263 |  | 1,630 | 633 |  |
| Primary outcome, n (%) | *Class not-OMI* | 8,242 (77.1) | 1,810 (80.0) | 0.003 | 1,390 (85.3) | 420 (66.4) | <0.001 |
|  | *Class OMI* | 2,450 (22.9) | 453 (20.0) | 0.003 | 240 (14.7) | 213 (33.6) | <0.001 |
| Unique ECGs, n |  | 18,616 | 3,254 |  | 2,016 | 1,238 |  |
| ECGs Outcome breakdown, n (%) | *MI ruled-out* |  | 1,866 (57.3) | 1.000 | 1,456 (72.2) | 410 (33.1) | <0.001 |
|  | *NOMI* |  | 469 (14.4) | 1.000 | 107 (5.3) | 362 (29.2) | <0.001 |
|  | *OMI* |  | 919 (28.2) | 1.000 | 453 (22.5) | 466 (37.6) | <0.001 |

Cat., category; EU, Europe; US, United States; SD, standard deviation; OMI, occlusion myocardial infarction; NOMI, non-occlusion myocardial infarction; ECG, electrocardiogram; MI, myocardial infarction.

### Test sets characteristics

Procedural characteristics of both testing cohorts are shown in Table 2. The overall test set included 3,254 ECGs from 2,263 unique contacts (20% OMI) of 2,222 patients (age 62±14 years, 67% males). Of these, 2,016 ECGs from 1,630 contacts (with 240 [14.7%] OMI) were from the internal EU testing cohort, and 1,238 ECGs from 633 contacts (with 213 [33.6%] OMI) were from the US testing cohort. The prevalence of OMI differed between the internal EU and the external US test set, 14.7% compared to 33.6%, respectively (p<0.001). Contacts included in the US test set were younger, had more ECGs recorded before catheterization and had more STEMI ECGs. Gender, peak troponin and the TIMI flow of culprit vessels did not differ significantly between the two cohorts.

**Table 2.** Procedural characteristics of the patient contacts in the EU and US test sets.

| Parameter | Cat. | Overall Test Sets (n=2,263) | Internal EU Test Set (n=1,630) | External US Test Set (n=633) | P-value |
| --- | --- | --- | --- | --- | --- |
| Pre-CAG ECG, n (%) | *STEMI* | 264 (11.7) | 154 (9.4) | 110 (17.4) | <0.001 |
|  | *not-STEMI* | 1999 (88.3) | 1476 (90.6) | 523 (82.6) | <0.001 |
| Average ECGs per patient, mean (SD) |  | 1.4 (0.9) | 1.2 (0.6) | 2.0 (1.2) | <0.001 |
| Admission Troponin T (ng/L), median [Q1,Q3] |  | 7.4 [4.2,13.5] | 7.4 [4.2,13.5] | NA | NA |
| Peak Troponin T (ng/L), median [Q1,Q3] |  | 31.8 [5.0,1457.2] | 11.8 [4.3,340.5] | 340.0 [11.0,2820.1] | <0.001 |
| CAG performed, n (%) |  | 1408 (62.2) | 948 (58.2) | 460 (72.7) | <0.001 |
| Time to CAG (hours), median [Q1,Q3] |  | 13.4 [2.5,19.6] | 17.3 [4.9,20.4] | 3.8 [0.8,15.1] | <0.001 |
| Time to CAG, n (%) | *Late (12-24h)* | 310 (22.1) | 129 (13.7) | 181 (39.3) | <0.001 |
|  | *Delayed (4-12h)* | 123 (8.8) | 72 (7.6) | 51 (11.1) | <0.001 |
|  | *Early (2-4h)* | 256 (18.2) | 167 (17.7) | 89 (19.3) | <0.001 |
|  | *Immediate (<2h)* | 715 (50.9) | 576 (61.0) | 139 (30.2) | <0.001 |
| Culprit vessel, n (%) | *None* | 1632 (72.1) | 1316 (80.7) | 316 (49.9) | <0.001 |
|  | *Native* | 605 (26.7) | 298 (18.3) | 307 (48.5) | <0.001 |
|  | *Graft* | 26 (1.1) | 16 (1.0) | 10 (1.6) | <0.001 |
| Culprit artery, n (%) | *LMCA* | 11 (1.7) | 6 (1.9) | 5 (1.6) | <0.001 |
|  | *LAD* | 234 (37.1) | 113 (36.0) | 121 (38.2) | <0.001 |
|  | *LCx* | 142 (22.5) | 59 (18.8) | 83 (26.2) | <0.001 |
|  | *RCA* | 220 (34.9) | 130 (41.4) | 90 (28.4) | <0.001 |
|  | *PDA* | 9 (1.4) | 0 (0.0) | 9 (2.8) | <0.001 |
|  | *RI* | 11 (1.7) | 2 (0.6) | 9 (2.8) | <0.001 |
|  | *Multi-vessel* | 4 (0.6) | 4 (1.3) | 0 (0.0) | <0.001 |
| Culprit stenosis (%), median [Q1,Q3] |  | 90.0 [70.0,100.0] | 80.0 [60.0,100.0] | 95.0 [90.0,100.0] | <0.001 |
| Culprit TIMI Flow, n (%) | *TIMI-0* | 244 (38.6) | 119 (37.8) | 125 (39.4) | 0.897 |
|  | *TIMI-1* | 38 (6.0) | 19 (6.0) | 19 (6.0) | 0.897 |
|  | *TIMI-2* | 70 (11.1) | 33 (10.5) | 37 (11.7) | 0.897 |
|  | *TIMI-3* | 280 (44.3) | 144 (45.7) | 136 (42.9) | 0.897 |
| Collateral flow, n (%) | *NONE* | 284 (90.4) | 284 (90.4) | NA | NA |
|  | *MILD* | 16 (5.1) | 16 (5.1) | NA | NA |
|  | *MODERATE* | 12 (3.8) | 12 (3.8) | NA | NA |
|  | *HIGH* | 2 (0.6) | 2 (0.6) | NA | NA |
| Time to Revascularization (hours), median [Q1,Q3] |  | 7.5 [2.1,19.3] | 7.5 [2.1,19.3] | NA | NA |
| Treatment, n (%) | *Conservative* | 706 (50.1) | 525 (55.4) | 181 (39.3) | <0.001 |
|  | *PCI* | 699 (49.6) | 422 (44.5) | 277 (60.2) | <0.001 |

Cat., category; EU, Europe; US, United States; CAG, coronary angiography; ECG, electrocardiogram; STEMI, ST-elevation myocardial infarction; SD, standard deviation; LMCA, left main coronary artery; LAD, left anterior descending artery; LCx, left circumflex artery; RCA, right coronary artery; PDA, posterior descending artery; RI, ramus interventricularis; TIMI, Thrombolysis In Myocardial Infarction; PCI, percutaneous coronary intervention.

### AI model performance

The OMI AI model with an optimal threshold (threshold of 0.1106) achieved an AUC of 0.941 (95% CI: 0.926, 0.954) in identifying the primary outcome of OMI (Panel A, Figure 2) on the overall test set. As shown in Figure 3, OMI AI model performance was comparable on both the European internal (Panel A) and US external testing datasets (Panel B), and achieved an AUC of 0.943 (95% CI: 0.925, 0.961) and of 0.918 (95% CI: 0.893, 0.942) respectively.

**Figure 2.**
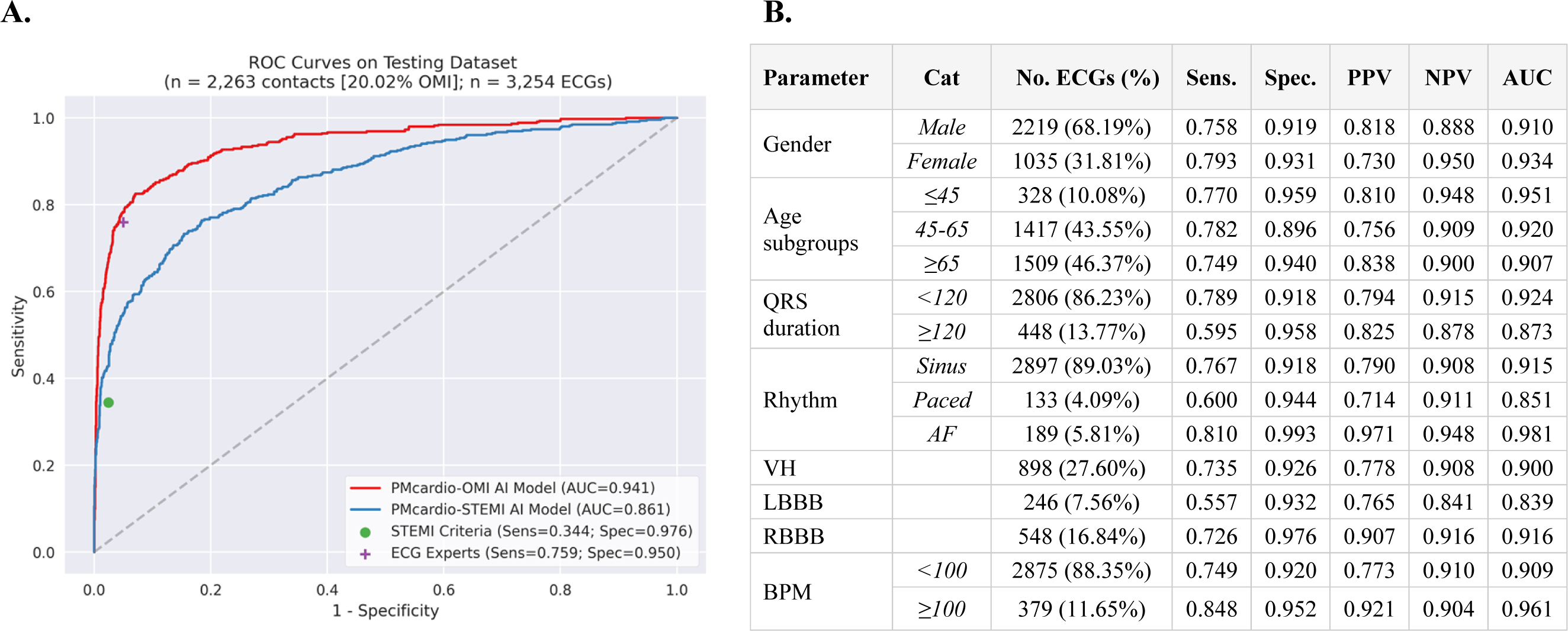
AI model performance on the overall test sets and subgroup analysis. *Panel A shows the ROC curve of AI algorithms, PMcardio-OMI (red) and PMcardio-STEMI (blue) and sensitivity and specificity of STEMI criteria (green dot) and ECG experts (purple cross) on combined EU and US testing cohorts. The AUC is 0.941 (n = 2,263 contacts [20.02% OMI]); Panel B shows the PMcardio-OMI AI model performance on different patient subgroups*. ROC, Receiver operating curve; OMI, Occlusion myocardial infarction; AI, artificial intelligence; STEMI, ST-elevation Myocardial Infarction; ECG, electrocardiogram; Sens, Sensitivity; Spec, Specificity; PPV, Positive predictive value; NPV, Negative predictive value; AUC, Area under the curve; AF, atrial fibrillation; VH, Ventricular hypertrophy; LBBB, Left bundle branch block; RBBB, Right bundle branch block; BPM, beats per minutes.

**Figure 3.**
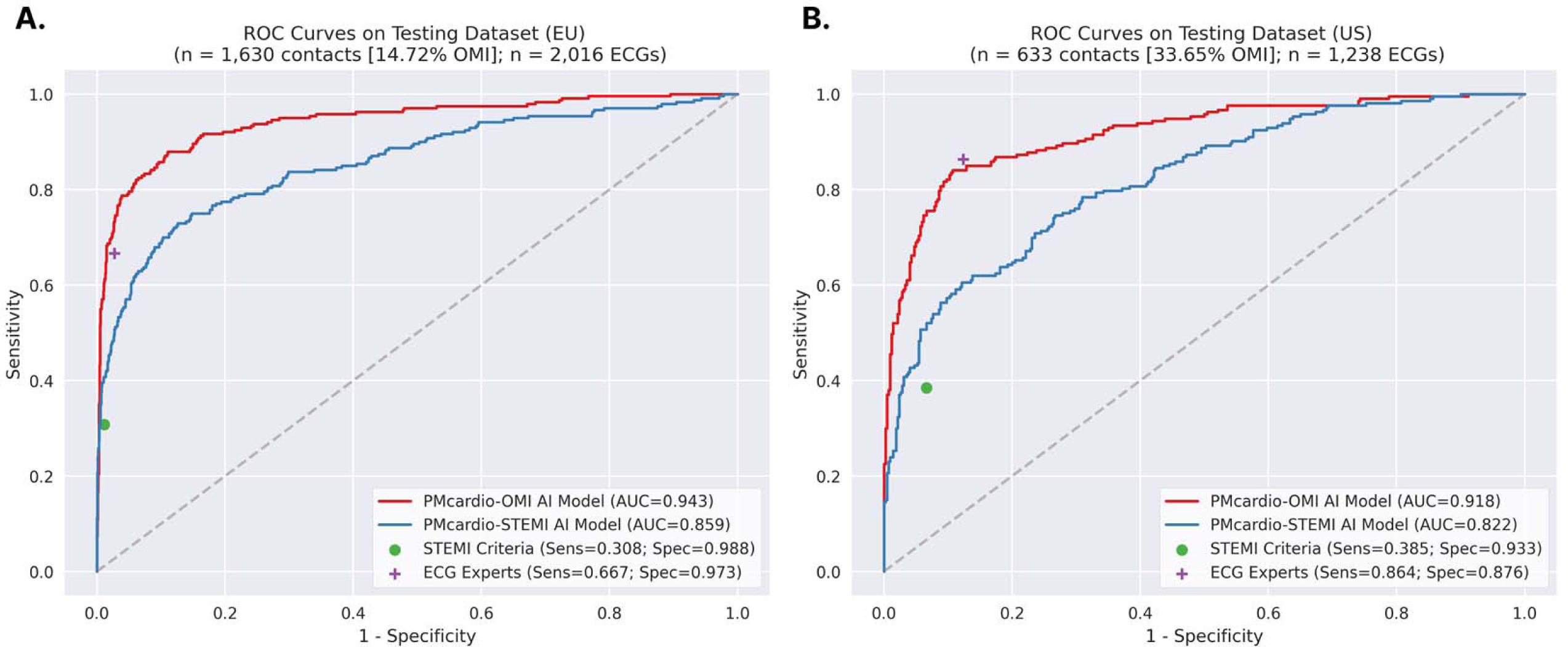
AI model performance on EU and US testing cohorts separated. *Figure shows the ROC curve of AI algorithms, PMcardio-OMI (red) and PMcardio-MI (blue) and sensitivity and specificity of STEMI criteria (green dot) and ECG experts (purple cross). Panel A shows the AUC of PMcardio-OMI AI of 0.943 (n=1,630 acts [14.72% OMI] on the EU internal testing cohort; Panel B shows the AUC of PMcardio-OMI AI of 0.918 (n=633 contacts [33.65% OMI] on the US external testing ort.* ROC, Receiver operating curve; EU, Europe; US, United States; OMI, Occlusion myocardial infarction; AI, artificial intelligence; STEMI, ST-elevation Myocardial Infarction; ECG, electrocardiogram; Sens, Sensitivity; Spec, Specificity; AUC, Area under the curve.

### Subgroup performance

AI model performance was tested across different subgroups and patient segments based on baseline characteristics and electrocardiographic patterns (Figure 2, Panel B). The model yielded stable performance across genders and age subgroups (ranging from 0.907 to 0.951 AUC). Significantly greater performance was recorded for ECGs with QRS duration under 120 milliseconds (0.924 [95% CI: 0.912, 0.935] AUC) and heart rate over 100 beats per minute (0.961 [95% CI: 0.940, 0.978] AUC), compared to their counterparts (0.873 [95% CI: 0.830, 0.914] and 0.909 [95% CI: 0.896, 0.922] AUC, respectively) (p<0.001). Model performance was comparable when tested on secondary definitions of OMI with different TIMI flow and troponin cut-off combinations, as well as the occurrence of PCI (Table 3).

**Table 3.** Performance of PMcardio-OMI AI models and analysis of different OMI outcome definitions across the grouped testing datasets (both EU and US). In bold primary outcome definition of OMI

| OMI outcome definition | PMcardio-OMI AI Model – Optimal threshold<sup>±</sup> |  |  |  |  |  |
| --- | --- | --- | --- | --- | --- | --- |
|  | *Sens.* | *Spec.* | *PPV* | *NPV* | *AUC* | *MCC* |
| Culprit TIMI 0-1 | 87.5%<br>(83.4-91.3) | 86.9%<br>(85.4-88.4) | 0.485<br>(0.445-0.528) | 0.980<br>(0.973-0.987) | 0.929<br>(0.912-0.944) | 0.588<br>(0.548-0.628) |
| Culprit TIMI 0-1 *OR* TIMI 2-3 Trop T ≥500 ng/L | 83.6%<br>(79.9-86.9) | 92.4%<br>(91.1-93.6) | 0.727<br>(0.688-0.765) | 0.959<br>(0.949-0.968) | 0.942<br>(0.928-0.955) | 0.722<br>(0.685-0.756) |
| Culprit TIMI 0-1 *OR* TIMI 2-3 with Trop T ≥1000 ng/L | 84.5%<br>(80.8-88.0) | 91.6%<br>(90.3-92.8) | 0.691<br>(0.65-0.73) | 0.964<br>(0.955-0.972) | 0.942<br>(0.928-0.955) | 0.706<br>(0.667-0.74) |
| Culprit TIMI 0-2 *OR* TIMI 3 with Trop T ≥500 ng/L | 81.9%<br>(78.1-85.3) | 93.1%<br>(91.9-94.2) | 0.754<br>(0.715-0.791) | 0.952<br>(0.942-0.962) | 0.939<br>(0.924-0.952) | 0.728<br>(0.69-0.762) |
| **Culprit TIMI 0-2 *OR* TIMI 3 with Trop T ≥1000ng/L** | **82.6%**<br>**(78.9-86.1)** | **92.8%**<br>**(91.5-93.9)** | **0.741**<br>**(0.7-0.778)** | **0.955**<br>**(0.945-0.965)** | **0.941**<br>**(0.926-0.954)** | **0.724**<br>**(0.687-0.758)** |
| Culprit TIMI 0-2 *OR* TIMI 3 with Trop T ≥1000 ng/L AND PCI performed | 82.4%<br>(78.6-86.0) | 92.6%<br>(91.3-93.7) | 0.733<br>(0.694-0.771) | 0.955<br>(0.945-0.965) | 0.939<br>(0.925-0.952) | 0.718<br>(0.68-0.753) |

± Optimal threshold based on ROC analysis (threshold of 0.1106)
OMI, Occlusion myocardial infarction; AI, artificial intelligence; STEMI, ST-elevation myocardial infarction; Sens., Sensitivity; Spec., Specificity; PPV, positive predictive value; NPV, Negative predictive value; AUC, Area under curve; MCC, Matthews correlation coefficient; TIMI, Thrombolysis In Myocardial Infarction; Trop, Troponin; PCI, percutaneous coronary intervention.

### AI model benchmarking

The OMI AI model was compared to three standard criteria assessing the same 12-lead ECGs in the overall test set for the presence of OMI (Table 4). In identifying OMI, the OMI AI model with optimal threshold recorded a significantly superior sensitivity of 82.6% (95% CI: 78.9%, 86.1%) compared to STEMI criteria and PMcardio-STEMI AI model [sensitivities 34.4% (95% CI: 30.0%, 38.8%) and 54.5% (95% CI: 49.7%, 58.8%), respectively], and statistically equal sensitivity compared to ECG experts [75.9% (95% CI: 71.9%, 80%)]. Accuracies were equal between PMcardio-OMI model and experts, and significantly higher than STEMI criteria and PMcardio-STEMI AI model. All benchmark criteria achieved high specificity on the overall test set ranging from 92.8% to 97.6%. Head-to-head comparison of confidence intervals of two evaluation criteria for OMI across all standard metrics is summarized in Supplemental Table 1. When adjudicated across six metrics (sensitivity, specificity, accuracy, PPV, NPV and MCC) the OMI AI Model showed statistically superior performance compared to STEMI criteria and PMcardio-STEMI AI model and equal (non-inferior) performance to ECG experts. ECG experts also recorded significantly better performance compared to STEMI criteria and PMcardio-STEMI AI model. Mean time to OMI diagnosis was significantly shorter for OMI AI model compared to STEMI criteria, 2.0 hours vs 4.9 hours respectively (p<0.001) (Figure 4), but comparable to ECG experts, with a mean time of 2.5 hours (p=0.12). Patients with OMI received interventions at a similar rate regardless of STEMI criteria presence and outcome definition (primary outcome definition, 96.5% vs. 94.5% [p=0.570]; strictest OMI outcome [TIMI 0-1 flow only], 96.3% vs. 92.4% [p=0.358]) (Supplemental Table 2).

**Figure 4.**
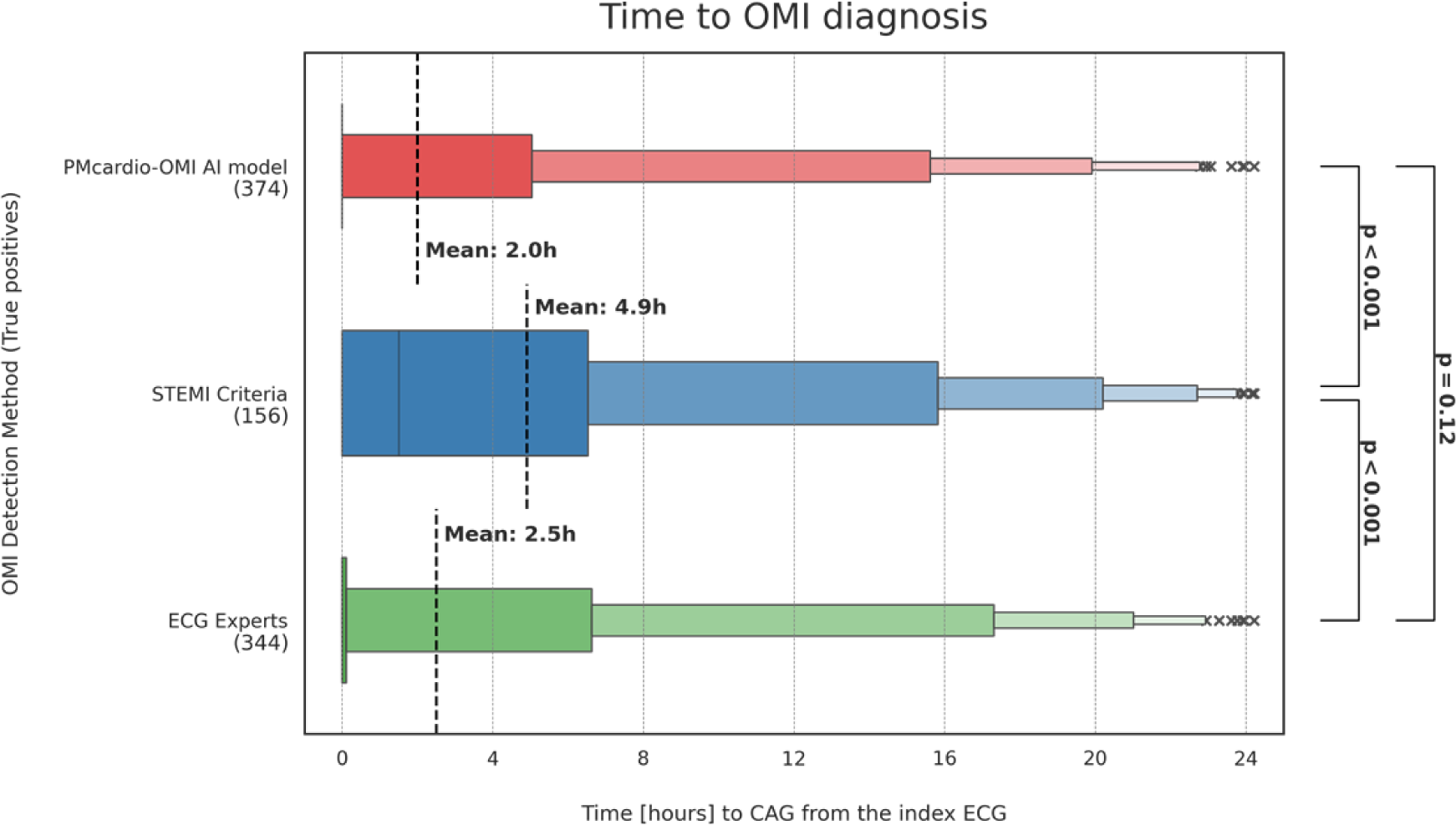
Time to OMI diagnosis by detection method. *Graph shows the time to OMI diagnoses (primary outcome definition) when relying on individual detection methods. If the detection method detected OMI on the first ECG, the time to diagnosis is 0. If the detection method did not catch OMI on any ECG, the time to diagnosis corresponds to the time to coronary angiography.*

**Table 4.**
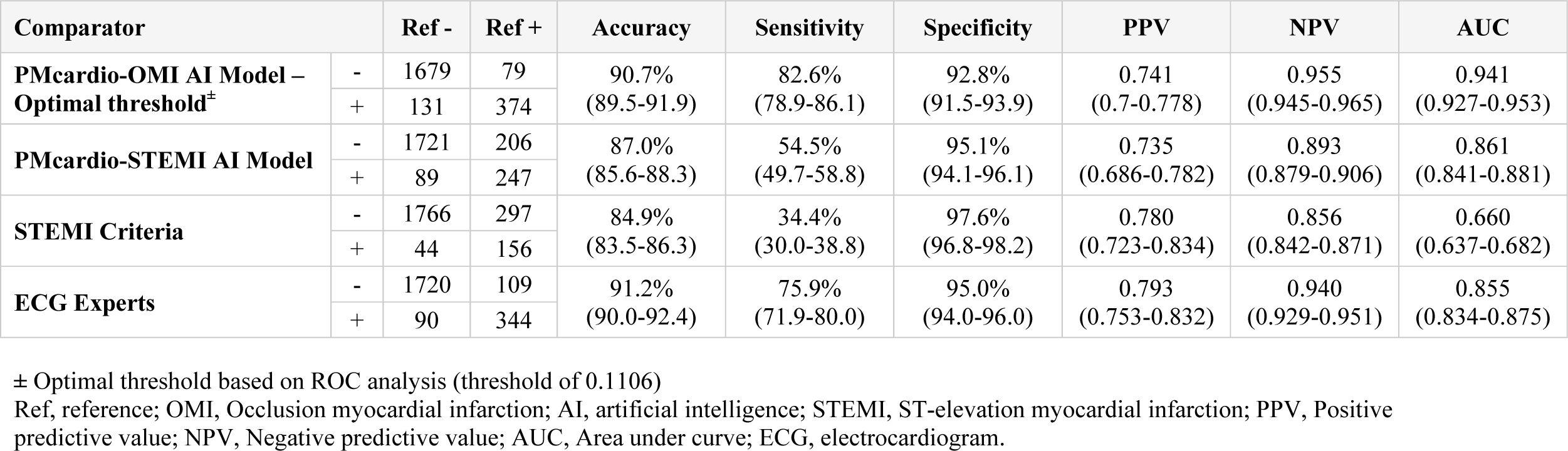
Head-to-head benchmark comparison in detecting the primary outcome definition of OMI.

### Analysis of misclassified cases

There were 131 subjects identified by the OMI AI model and 44 subjects identified by STEMI criteria as OMI on ECG who did not meet the primary outcome definition of OMI, for purpose of the manuscript termed as “false positives” (Supplemental Table 3). However, false positives of the AI model and STEMI criteria significantly differed in the rate of AMI and AMI-related interventions. Of the 131 AI model false positives, 71 (54.2%) had AMI, 45 (34.4%) had acute culprit lesions, and 40 (30.5%) had AMI with PCI. In contrast, only 15 (34.1%) (p=0.035) of STEMI criteria false positives had AMI, 6 (13.6%) had acute culprit lesions (p=0.031), and 5 (11.4%) had AMI with PCI (p=0.036). In other words, false positives by STEMI criteria were more often cases without any AMI (whether OMI or NOMI), whereas with AI, they were much more often actual AMI cases that needed intervention, albeit not necessarily emergently. Of the 297 OMI patients (65.5% of all OMI) missed by STEMI criteria, only 100 (33.6%) had a time to revascularization of under 2 hours. Of the remaining 197 OMI patients without positive STEMI criteria who did not get urgent CAG, 112 (56.8%) were correctly identified by the OMI AI model using the first ECG; these patients had a median revascularization time 10.9 hours (IQR 5.3,17.3).

## DISCUSSION

In this study, we validated the novel PMcardio-OMI AI model which is comparable to specialized OMI ECG experts in detecting invasively confirmed acute coronary occlusion from individual 12-lead electrocardiograms recorded in ACS patients before cardiac catheterization, blinded to all other clinical information. High accuracy was upheld across two large, independent testing cohorts of ACS patients from Europe and United States, with robust performance across all subgroups and on ECGs recorded at various time intervals before coronary angiography.

The present research is driven by the unmet need related to the suboptimal triage of ACS patients at their presentation. Barely 25% of patients with ACS present with typical ST-segment elevation on their initial ECG^30^ and up to 35% of patients without such ST-segment elevation have total coronary occlusion on the delayed angiography.^31-35^ In addition, 20% of OMI met STEMI criteria on the initial ECG, 30% on serial ECGs, and only 49% were recognized by cardiologists as STEMI.^36,37^ Compared to NSTEMI with a non-occlusive stenosis of the culprit coronary artery (NOMI)^21^, patients with OMI have far higher mortality and worse left ventricular function, in spite of having younger age and fewer comorbidities.^4^

Several previous studies deployed machine learning to triage patients presenting with ACS, however, bearing multiple limitations.^30,38-50^ The majority of these studies did not validate the occlusive or flow-limiting culprit lesions on coronary angiogram and relied on a subjective majority vote of board-certified cardiologists interpreting the ECG with STEMI as the surrogate for OMI.^30,39-43^ They often employed a spectrum of input clinical features in addition to the ECG waveform restricting their practical, real-world implementation.^44-50^ Finally, their validation was not scrutinized in sizeable external and international datasets or on ECGs recorded from multiple ECG device vendors limiting their applicability to digital ECG file formats from a single manufacturer.

Our study is characterized by several methodological strengths. First, it is built using an international cohort of standardized 12-lead ECG waveforms of different formats from multiple vendors, either paper form or screenshot images. Second, the ACO reference standard used for model evaluation was an objective, invasively collected composite of TIMI flows of culprit vessels and biomarker elevation. Using this robust methodology, the OMI AI model yielded superior accuracy in the validation within an independent cohort. Likewise, the AI model demonstrated sustained high performance (>0.92 AUC) on both EU internal testing datasets with the natural prevalence of OMI within a cohort of ACS patients and an external validation set of patients from two independent US centers. The OMI AI model yielded a statistically superior performance to STEMI criteria and equal performance to ECG experts when compared using six complementary performance metrics. More specifically, the model outperformed the standard ECG millimeter criteria in detecting ACO and provided an over twofold increase in sensitivity while maintaining the high specificity in STEMI criteria. The presented OMI AI model detects OMI significantly earlier (by 2.9 hours) compared to current guideline recommended STEMI criteria. Finally, the false positive OMI interpretations classified by the AI model were more likely to have acute MI (though not meeting primary outcome criteria), have an acute culprit lesion, and undergo PCI, compared to the false positive interpretations by STEMI criteria.

### Clinical implications

Our study has several implications for the future management of ACS. OMI AI model paired with proprietary digitization technology offers accurate detection of patients with ACO with occlusive or flow limiting lesion using a single 12-lead ECG tracings independent of ECG vendor or its format. Specifically, such accurate and timely ECG-based ACS diagnosis at the time of first patient contact could prompt swift coronary intervention as recommended currently in case of standard STEMI criteria. The rapid reperfusion in such management can consequently limit the burden of myocardial injury with favorable impact on clinical outcomes. In this regard, the model reliably detected ACO on average 2.9 hours before the current guideline-based ECG standards suggesting its potential to streamline the timely referral of ACS patients at risk for poor outcomes.

### Limitations

Several limitations are to be considered. Although validated on multi-center, international cohorts of patients, our study lacks prospective validation. In clinical practice, the decision to refer for early angiography in patients presenting with NSTEMI is not only based on the ECG but encompasses often additional criteria. Nevertheless, our results show nearly half (45.4%) of NSTEMI-OMI patients that could have had accelerated access to PCI based on the AI model detection truly underwent revascularization within 2 hours. However, their median time to revascularization was delayed over 10 hours. Annotation of STEMI criteria may be subjective, and we have only included one interpretation per ECG for this metric. Although the model has demonstrated robust performance across various patient subgroups, our study lacks AI model explainability. The OMI AI model detects OMI with a binary granularity. It is understood that the different stages of culprit coronary lesion leading to acute coronary syndrome, in terms of dynamics (active or reperfused) and time (acute or subacute), can have an influence on patient outcomes and the timing of invasive strategies. Lastly, our study was not designed to quantify other relevant clinical endpoints such as mortality, in-hospital complications, or MACE. Future work should address these limitations and observe the AI model efficacy and clinical benefit deployed in a prospective cohort of ACS patients.

### Conclusions

We have developed and validated PMcardio-OMI AI model able to accurately detect ACS patients with angiographically confirmed occlusion of culprit coronary arteries using only single standard 12-lead ECGs in a large international, multi-center cohort of ACS patients. Our AI model outperformed gold-standard STEMI criteria in the diagnosis of OMI and warrants further prospective clinical studies to define the role of OMI AI model in guiding ACS triage and timely referral of patients benefiting from immediate revascularization.

## ONLINE CONTENT

The dataset is not available for public sharing, given our institutional review board approval restrictions. The OMI AI ECG model is available for external validation, benchmarking and research use at: https://bit.ly/omi-ai-ecg.

### Disclosures

Dr. Herman is the Co-founder and Chief Medical Officer of Powerful Medical; Michal Martonak, Jakub Bahyl, Andrej Iring, Boris Vavrik, Vladimir Boza, Viera Kresnakova and Anthony Demolder are employees and shareholders of Powerful Medical. Dr. Smith, Dr. Meyers and Dr. Perl are shareholders in Powerful Medical. Dr. Herman, Dr. Bertolone, Dr. Leone, Dr. Viscusi are supported by a research grant from the CardioPaTh PhD Program. Other authors report no conflict of interest.

## Data Availability

The OMI AI ECG model is available for external validation, benchmarking and research use at: https://bit.ly/omi-ai-ecg. The full dataset is not available for public sharing, given our institutional review board approval restrictions.

## Acknowledgements

Acknowledgements:

The authors would like to express the appreciation to the clinical experts, study team, data scientists and AI engineers supporting the data collection, processing, and validation.

